# Estimating Chronic Kidney Disease Stage Transitions from Irregular Electronic Health Record Data Using an Expectation–Maximization Framework

**DOI:** 10.64898/2026.03.08.26347890

**Authors:** Wendy Qi, Jennifer M. Lobo, Guofen Yan, Rahwa Ghenbot, Kenneth G. Sands, Tracey L. Krupski, Stephen H. Culp, Daniel Otero-León

**Affiliations:** University of Virginia School of Engineering; University of Virginia School of Medicine

## Abstract

**Objective:** To estimate chronic kidney disease (CKD) stage transition probabilities in patients with small renal masses (SRMs) using irregularly observed electronic health record (EHR) data, addressing challenges of interval censoring and irregular measurement intervals in real-world clinical practice.

**Data Sources:** We used EHR data from the University of Virginia Small Renal Mass (SRM) registry (2006–January 2026), capturing outpatient renal function data prior to any definitive treatment. CKD stages were defined using estimated glomerular filtration rate (eGFR) thresholds based on KDIGO guidelines.

**Study Design:** The final analytic cohort included 527 patients with at least two outpatient eGFR measurements prior to definitive treatment. We applied an expectation–maximization (EM) algorithm to estimate discrete-time CKD stage transition matrices while accounting for irregular follow-up and unobserved intermediate transitions. Transition matrices were estimated under 3-and 6-month cycle lengths overall as well as stratified by age and sex. Likelihood ratio tests were used to compare EM-based estimates with a naïve one-step counting estimator.

**Results:** The EM framework yielded clinically plausible transition structures dominated by self-transitions and progression primarily to adjacent CKD stages, with reduced spurious backward transitions relative to the naïve estimator. Transition patterns were consistent across 3- and 6-month cycle lengths. Age-stratified analyses showed that older patients had slightly higher probabilities of progression to more advanced CKD stages compared with younger patients, whereas sex-stratified differences were minimal. Likelihood ratio comparisons supported the consistency of the EM-based models with the observed transition data in both the overall cohort and subgroup analyses.

**Conclusions:** The EM approach provides a principled and computationally efficient method for estimating CKD stage progression from irregularly observed EHR data, yielding transition matrices suitable for discrete-time decision-analytic and health economic models.

## 1. Introduction

### 1.1 Background

Chronic kidney disease (CKD) is a progressive condition characterized by the gradual loss of kidney function over time. It affects more than 800 million people worldwide and is associated with high morbidity, mortality, and healthcare burden.^1,2^ An estimated 10–15% of adults have CKD, making it a major public health challenge due to its strong association with cardiovascular disease, end-stage kidney disease (ESKD), hospitalization, and premature death.^3–5^ Accurate characterization of CKD progression is therefore critical for predicting clinical outcomes and informing population-level planning.

CKD is classified into five stages based on estimated glomerular filtration rate (eGFR), ranging from stage 1 (eGFR ≥90 mL/min/1.73m²) to stage 5 (eGFR <15 mL/min/1.73m² or dialysis).^6^

Disease progression across CKD stages is heterogeneous, with some patients remaining stable for years while others progress rapidly to ESKD.^5,7^ In decision-analytic models, this progression is represented using stage-to-stage transition probabilities, which quantify the likelihood of moving between CKD stages over a given time horizon and serve as core inputs to Markov cohort models, patient-level microsimulations, and Markov decision processes (MDPs) used to project long-term outcomes and evaluate interventions.^8–10^

Estimating these transition probabilities relies on longitudinal kidney function data, typically obtained from repeated eGFR measurements in electronic health record (EHR) systems.

However, such data are collected at irregular and patient-specific time points and frequently contain missing measurements or censoring due to death, transplantation, or loss to follow-up. Many studies exclude patients with missing measurements or irregular visit patterns, introducing selection bias into transition estimates.^5,11^ Others apply simple Markov models without accounting for unobserved stage changes between visits.^12^ These limitations prevent accurate modeling of CKD progression in large EHR datasets, leading to unreliable projections and suboptimal guidance for clinical and policy decision-making.

In this study, we focus on CKD progression within a clinically important subpopulation: patients with small renal masses (SRMs). Prior institutional cohorts have reported that approximately 22% of patients with solid renal masses present with CKD stage 3 or higher at baseline.^13^ Beyond this substantial baseline burden, patients with SRMs are at elevated risk for further renal function decline due to older age, high comorbidity burden, and potential nephron loss associated with treatment-related effects.^13–15^ Preservation of kidney function is therefore a central consideration in SRM management, alongside cancer control.^16–19^ Importantly, CKD progression patterns in patients with SRMs may differ from those observed in broader CKD populations, and transition estimates derived from general CKD cohorts may not adequately reflect the clinical dynamics of this group. Moreover, follow-up in SRM management is inherently irregular and driven by surveillance schedules and treatment decisions, further motivating the need for methods that explicitly accommodate irregularly collected data. Reliable characterization of CKD progression in this population is essential for evaluating trade-offs between active surveillance and intervention strategies and for supporting personalized treatment decision-making.

To address this need, we apply an Expectation–Maximization (EM) algorithm to estimate CKD stage transition probabilities from longitudinal EHR data in a cohort of patients with SRMs managed under active surveillance. The objectives of this study are to: (1) estimate CKD stage-specific transition probability matrices in patients with SRMs that explicitly account for irregular observation intervals and unobserved intermediate transitions; (2) quantify differences between EM-based estimates and conventional naïve counting approaches in terms of transition probabilities and model fit; and (3) provide empirically derived, model-ready CKD transition probability matrices to support future state-transition modeling and decision-analytic studies.

Our work highlights the importance of appropriately handling irregularly observed CKD data in patients with SRMs and establishes a methodological foundation for more accurate estimation of stage-specific transition probabilities in this population, while providing a framework that can be extended to other CKD populations with irregular follow-up.

### 1.2 Related Work

A large body of literature has examined methods for estimating disease progression from longitudinal clinical data. These methods span multiple statistical and modeling paradigms and differ in how disease states, transition timing, and observation processes are represented. In this section, we organize prior work into several major methodological families, including naïve counting and empirical approaches, discrete-time and continuous-time Markov models, survival and hazard-based models, hidden Markov and latent-state models, and expectation–maximization (EM) and likelihood-based approaches for interval-censored multistate processes. For each family, we briefly summarize core ideas and typical applications, with emphasis on relevance to modeling chronic kidney disease stage transitions.

#### Naïve counting estimators

The simplest approaches estimate disease progression by directly counting observed transitions between consecutive measurements and normalizing by the person-time at risk in each origin state. These estimators are straightforward to implement and require minimal modeling assumptions. However, they rely solely on observed transitions and therefore do not account for interval censoring or unobserved intermediate state changes occurring between clinical visits. In settings with irregular or infrequent observations, such as EHR-derived longitudinal data, this limitation can lead to biased estimates, typically underestimating true progression rates.^12,20^

#### Survival and Hazard Models

Survival and hazard-based models treat disease progression as a time-to-event process and are the most commonly used approach in CKD progression research, particularly for modeling time to ESKD and competing risk of death. Multi-state survival models, including illness-death formulations, extend standard survival analysis to multiple disease states. Boucquemont et al. showed that illness-death models provide more accurate estimates of CKD progression hazards than standard survival models when progression times are interval-censored and death is a competing risk.^21^ Parametric survival and accelerated failure time models have also been proposed to analyze CKD progression using surrogate endpoints based on eGFR decline or slope-based thresholds.^22^

Because kidney function is both a longitudinal biomarker and a predictor of progression, several studies have adopted joint longitudinal-survival models that link eGFR trajectories to time-to-event outcomes. Van den Brand et al. demonstrated that joint models outperform Cox models using baseline or slope-based eGFR for predicting kidney failure, and Liao et al. extended this framework to personalized dynamic assessment of CKD progression.^23,24^ As an alternative strategy, landmark and dynamic prediction models update risk using the most recent biomarker values, but they typically assume that measurement times occur independently of the underlying disease process.^25^

Overall, survival and hazard models provide direct inference on event risks and are supported by mature software, but they estimate continuous-time hazards rather than full stage-to-stage transition matrices. Moreover, standard implementations can be biased when interval censoring is ignored and typically do not model the observation process, limiting their direct applicability for estimating comprehensive disease progression dynamics from irregular EHR data.

#### Continuous-Time Markov Models (CTMCs)

Continuous-time Markov models represent disease progression as a continuous-time stochastic process governed by transition intensity (Q) matrices that characterize instantaneous rates of movement between states. Unlike discrete-time models, CTMCs naturally accommodate irregular observation schedules by linking intensities to transition probabilities through matrix exponentials, thereby avoiding arbitrary discretization.

In CKD, Begun et al. developed a six-state nonhomogeneous CTMC including CKD stages 3–5, dialysis, kidney transplant, and death, with covariate effects on transition intensities specified through proportional hazards-type modifiers.^26^ Their model was fit using maximum likelihood to irregularly observed clinical data and enabled personalized prediction of progression trajectories. More generally, Jackson provides a comprehensive likelihood-based framework for fitting CTMCs to panel data, including both time-homogeneous and time-inhomogeneous models with covariates,.^27^ Kendall et al. showed that violations of the time-homogeneity assumption can lead to biased estimates and advocated for time-inhomogeneous formulations when supported by the data.^28^

Overall, CTMCs provide a principled framework for estimating stage-to-stage transition dynamics under irregular observation. However, they can be computationally demanding for large state spaces or complex covariate structures. In standard formulations, CTMCs assume exponentially distributed holding times and often time-homogeneous transition intensities, which may be unrealistic for chronic diseases such as CKD where progression risk can vary over time.

#### Hidden Markov and Latent-State Models

Hidden Markov models (HMMs) and related latent-state formulations provide a natural framework for disease progression when true disease states are imperfectly observed due to measurement error or misclassification. In CKD, Luo et al. applied HMMs to stage transitions derived from EHR data, allowing observed CKD stages to be misclassified because of variability in eGFR measurements.^29^ Using discretized time windows (30, 90, and 180 days) and EM-based forward-backward estimation, they demonstrated through simulation that HMMs can reduce bias relative to naïve counting when measurement error is present, although discretization still introduces timing bias under irregular follow-up. Hu et al. proposed a nonparametric multistate approach that treats the underlying kidney function trajectory as a latent process observed with error, using penalized splines to smooth longitudinal eGFR measurements and derive transition probabilities from the smoothed latent trajectory.^30^ More generally, Bureau et al. developed continuous-time HMMs for misclassified disease outcomes with EM-based estimation of transition intensities and misclassification probabilities, allowing for asymmetric false-positive and false-negative rates.^31^

A key limitation of standard HMMs is the assumption that observation times are independent of the latent disease state. In reality, patients with more severe disease are often monitored more frequently. To relax this, Alaa and van der Schaar introduced hidden absorbing semi-Markov models which accommodate irregularly sampled and informatively censored data.^32^ Furthermore, Bartolucci and Farcomeni utilized shared-parameter models to link latent disease states directly to both longitudinal outcomes and dropout risks.^33^ For CKD specifically, Lu et al. developed joint latent-random-effects models that simultaneously account for longitudinal biomarkers, dependent observation times, and competing terminal events.^34^

Despite their conceptual elegance, HMMs face significant hurdles in large-scale EHR applications. These models are computationally intensive, require high-dimensional integration, and often suffer from identifiability issues, where the model cannot distinguish between a high rate of disease progression and a high rate of measurement error. Such challenges can limit their scalability and practical implementation in clinical decision-support settings.

#### Expectation-maximization (EM) algorithms for Interval-Censored Multi-State Models

The EM algorithm^35^ provides a principled framework for maximum likelihood estimation in the presence of incomplete, censored, or irregularly spaced data. For multi-state models with interval censoring, the EM approach treats unobserved intermediate transitions as missing data and iteratively computes expected transition counts (E-step) and updates transition probability estimates (M-step) until convergence.^36–38^ EM treats unobserved CKD stage transitions as latent variables and iteratively computes the expected number of transitions and the expected time spent in each stage, given the observed endpoints. By integrating over all possible latent paths between measurements, EM accommodates irregular visit timing, avoids excluding patients with sparse data, and yields unbiased transition probability estimates. This approach has been applied to various disease progression contexts but has seen limited use in CKD progression modeling despite its theoretical advantages for EHR data.^37,39–41^ This makes it particularly suitable for large retrospective CKD cohorts in which follow-up patterns vary widely across patients.

Within this class of methods, Sherlaw-Johnson et al. introduced an EM algorithm for estimating discrete-time Markov transition matrices from panel data with irregular observation intervals.^42^ This method reconstructs expected one-step transitions between states that are consistent with observed endpoints and updates the transition matrix by normalizing these expected counts, directly yielding discrete-time transition probabilities without assuming an underlying continuous-time process. Related methodological developments include composite or pseudo-likelihood approaches that trade statistical efficiency for computational simplicity, imputation- or simulation-based EM methods that use Monte Carlo sampling to generate latent paths, and formulations that allow transition probabilities to vary with time or patient covariates.

Relative to other EM applications, EM for discrete-time Markov models targets discrete-time transition probabilities and assumes disease states are observed without error, whereas EM for CTMC estimates intensity matrices and EM for HMM focuses on latent states and misclassification. The discrete-time EM framework therefore occupies a middle ground between CTMC and HMM approaches. Despite these advantages, EM-based discrete-time Markov methods have seen limited application in CKD progression research, where survival and CTMC remain dominant.

Taken together, existing approaches highlight important tradeoffs in CKD progression modeling. Survival and hazard models provide robust inference for specific event times but do not directly yield full stage-to-stage transition structures. CTMCs offer a principled continuous-time representation but can be computationally demanding and often rely on restrictive assumptions such as time-homogeneous intensities. HMMs and latent-state models accommodate measurement error and misclassification but introduce substantial computational and identifiability challenges. These limitations motivate a complementary and comparatively simple class of methods that estimate discrete-time Markov transition matrices from irregularly observed EHR data using EM. In particular, the approach of Sherlaw-Johnson et al. reconstructs expected one-step transitions under interval censoring to estimate stage-to-stage transition probabilities. The next section describes our implementation of this EM-based discrete-time Markov framework for CKD progression among SRM patients using EHR data.

## 2. Methods

### 2.1 Data Source and Cohort Description

We conducted a retrospective cohort study using EHR data from the University of Virginia (UVA) SRM registry, which contains demographic, laboratory, imaging, and treatment information for patients evaluated for small renal masses at UVA Health. The institutional review board–approved SRM registry contains retrospectively collected data from 2006 to 2015 and prospectively collected data since 2015. The current analysis includes data through January 2026.

The cohort was constructed to support estimation of CKD progression using the EM framework. Because the objective is to estimate CKD stage transitions prior to treatment, regardless of whether or when a patient eventually undergoes treatment, we included both patients managed without treatment and patients who later received treatment, using only renal function measurements obtained prior to treatment for the latter group. Renal function observations obtained during inpatient care were not included. Patients were included if they met the following criteria: (1) diagnosis of a small renal mass and inclusion in the UVA SRM registry; (2) a valid SRM conference date; (3) availability of eligible outpatient renal function measurements within a two-year lookback window preceding the SRM diagnosis date or later; and (4) at least two valid outpatient eGFR measurements, defined as two measurements during follow-up for patients without treatment and two pre-treatment measurements for patients who later underwent treatment. Patients not meeting these criteria were excluded. The cohort inclusion and exclusion process is summarized in Figure 1.

**Figure 1.**
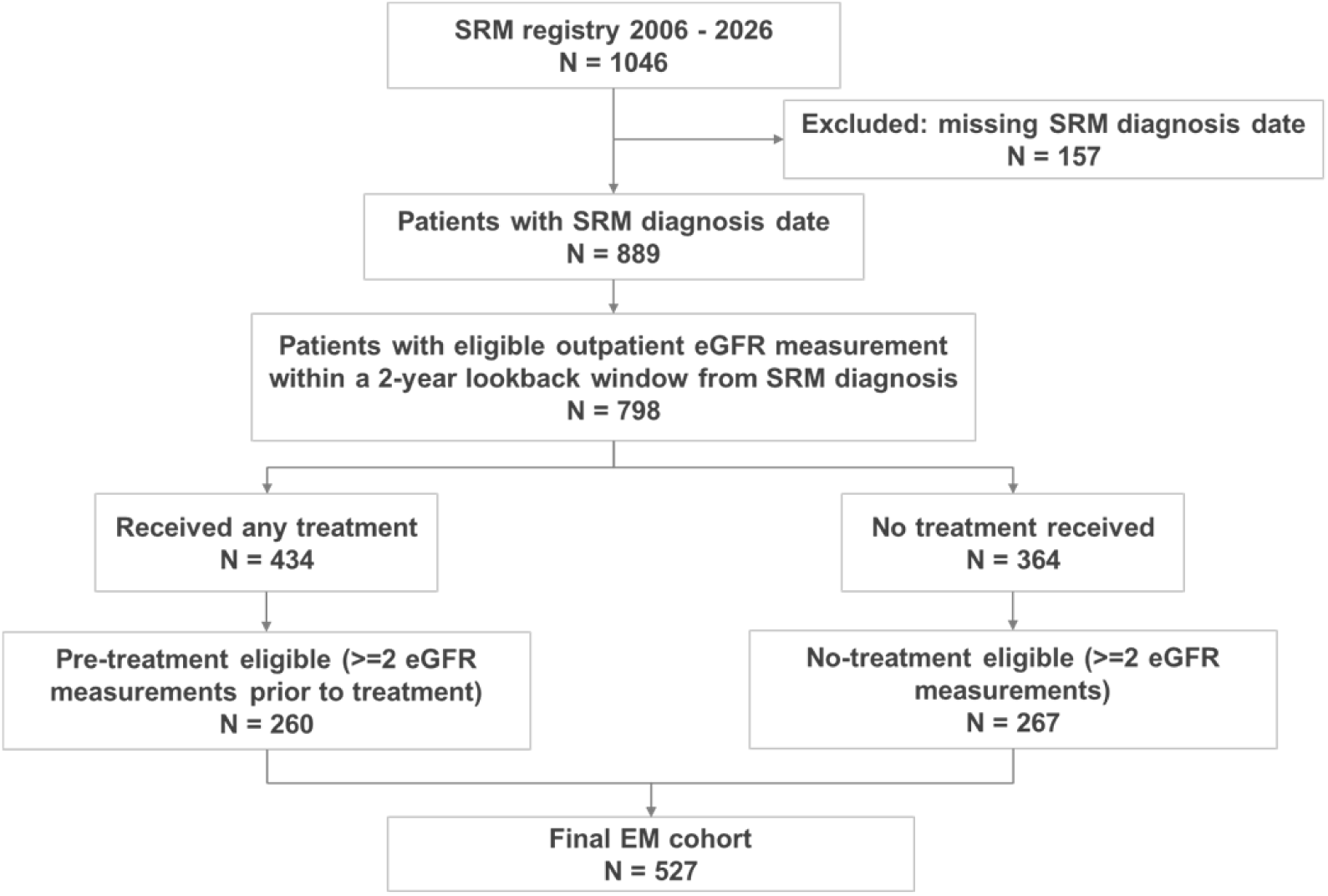
Cohort Selection for EM-based CKD Progression Modeling

We identified 1,046 patients in the SRM registry. After excluding patients without a valid SRM diagnosis date, 889 patients remained. Restricting outpatient eGFR measurements to those obtained within a two-year lookback window preceding diagnosis or later yielded 798 patients with eligible renal function data. These patients were stratified by treatment status, with 364 untreated and 434 treated patients. After requiring at least two valid outpatient eGFR measurements restricted to the pre-treatment period for treated patients, the final analytic cohort for EM-based CKD transition modeling consisted of 527 patients, including 267 in the no-treatment arm and 260 in the treated arm.

Baseline was defined in an arm-specific manner. For patients in the no-treatment arm, the observation window began two years prior to the SRM diagnosis date, and baseline was defined as the date of the first eligible outpatient eGFR measurement within this window. Follow-up extended through the study end date (January 2026). For patients in the treated arm, the observation window similarly began two years prior to the SRM diagnosis date, but renal function measurements were further restricted to those occurring prior to treatment; baseline was defined as the date of the first eligible pre-treatment outpatient eGFR measurement, and follow-up ended at the last outpatient eGFR measurement occurring before the first treatment date. eGFR was calculated from serum creatinine using the 2021 CKD-EPI Creatinine Equation,^43^ and CKD stage was determined by the eGFR value (Table 2). Measurements of eGFR taken during inpatient care were excluded in CKD progression estimation due to their transient nature obscuring the overall evolution of a patient’s kidney function. In this study, CKD 1 includes patients with normal kidney function and does not necessarily indicate kidney disease. Baseline characteristics included age, sex, race/ethnicity, diabetes mellitus, and baseline CKD stage.

**Table 1:** Baseline Characteristics of the Study Cohort

| Characteristic | Combined (N=527) |
| --- | --- |
| Number of patients | 527 |
| Age at diagnosis, mean (SD) | 62.6 (13.0) |
| Age at diagnosis, median [IQR] | 64.0 [55.0, 72.0] |
| **Sex** |  |
| M | 298 (56.5%) |
| F | 229 (43.5%) |
| **Race** |  |
| White/Caucasian | 426 (80.8%) |
| Black/African American | 80 (15.2%) |
| Asian | 3 (0.6%) |
| Other | 12 (2.3%) |
| Unknown | 6 (1.1%) |
| **Diabetes** | 182 (34.5%) |
| **Baseline CKD stage** |  |
| CKD 1 | 171 (32.4%) |
| CKD 2 | 217 (41.2%) |
| CKD 3a | 73 (13.9%) |
| CKD 3b | 33 (6.3%) |
| CKD 4 | 17 (3.2%) |
| CKD 5 | 16 (3.0%) |

**Table 2:** CKD Staging Based on eGFR thresholds

| GFR-based Stage | eGFR (mL/min/1.73 m<sup>2</sup>) | Description of kidney function |
| --- | --- | --- |
| **1** | ≥ 90 | Normal or high |
| **2** | 60–89 | Mildly decreased |
| **3a** | 45–59 | Mildly to moderately decreased |
| **3b** | 30–44 | Moderately to severely decreased |
| **4** | 15–29 | Severely decreased |
| **5** | < 15 | Kidney failure |

Cohort summary statistics are presented in Table 1. The mean age at diagnosis was 62.6 years (SD 13.0), and the cohort was relatively balanced by sex, with 56.5% male patients. The cohort was predominantly White/Caucasian (80.8%), with 15.2% Black/African American patients, and approximately one-third of patients had diabetes mellitus (34.5%). At baseline, most patients had preserved kidney function, with 73.6% in CKD stages 1–2 and 26.4% in CKD stage 3 or higher. Given the substantial heterogeneity in clinical follow-up, both mean and median summaries are reported to account for skewed distributions. The median follow-up duration was 0.5 years (IQR: 0.0–1.4; mean: 1.1, SD: 1.7). Patients contributed a median of 2 outpatient eGFR measurements (IQR: 1.0–4.0; mean: 5.4, SD: 10.3), with a median inter-measurement interval of 1.2 months (IQR: 0.3–4.1; mean: 3.1, SD: 4.5).

### 2.2 Transition Probability Estimation Using the EM Algorithm

#### 2.2.1 Markov Model and Likelihood

We modeled kidney function progression using a discrete-time Markov model, in which patients transition between CKD stages over time. Under the Markov assumption, the probability of transitioning to a future CKD stage depends only on the current stage and not on the full history of prior stages. Although CKD progression may exhibit path dependence in practice (e.g., rates of decline may vary with prior trajectory), the Markov assumption provides a standard and pragmatic approximation that has been widely adopted in decision-analytic models.

The model state space consists of six CKD stages corresponding to KDIGO guideline definitions based on eGFR thresholds. Death was not included as a model state, as the objective was to estimate CKD stage progression conditional on survival, with mortality incorporated separately in downstream decision-analytic models. Let 𝑆(𝑡) ∈ {1, 2, 3𝑎, 3𝑏, 4, 5} denote the CKD stage at time 𝑡. Let 𝑃 denote the one-step transition probability matrix, where each element 𝑃_𝑖𝑗_ denote the probability of transitioning from CKD stage 𝑖 to 𝑗 over one unit of time:

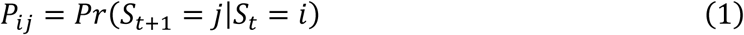

where 𝑃_𝑖𝑗_ ≥ 0 and ∑_𝑗_ 𝑃_𝑖𝑗_ = 1 for all states 𝑖.

If a transition is observed from stage 𝑖 to 𝑗 after 𝑡 time units, the probability of this observation is given by the (𝑖, 𝑗) element of the matrix 𝑃^𝑡^, denoted (𝑃^𝑡^)_𝑖𝑗_. Let 𝑂_𝑖𝑗𝑡_ denote the number of observed transitions from CKD stage 𝑖 to stage 𝑗 occurring over 𝑡 time units across all patients. The likelihood of the observed data 𝑌, given transition matrix 𝑃, is then:

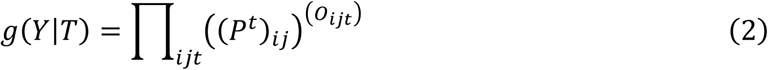

Taking logarithms yields the log-likelihood:

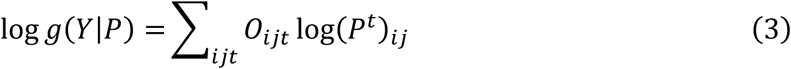

Direct maximization of the log-likelihood function is difficult because it involves summation over all possible hidden pathways, creating a nonlinear and non-convex optimization problem. We therefore estimate transition matrix 𝑃 using the EM algorithm which addresses this optimization challenge by introducing latent variables representing unobserved intermediate transitions and iteratively maximizing an expected complete-data log-likelihood that is easier to optimize.

#### 2.2.2 EM Algorithm Formulation

To estimate the transition matrix 𝑃, we followed the approach of Sherlaw-Johnson et al.^42^ for Markov processes with incomplete observation. In our context, the EM algorithm treats the unobserved CKD transitions occurring between observation times as latent data. The algorithm alternates between E- and M-steps.

In the E-step, the algorithm computes the expected number of one-step transitions between CKD stages, conditional on the observed data and the current estimate of the transition matrix.

Formally, it computes 𝑆_𝑖𝑗_(𝑃^(𝑝)^) = 𝐸[𝑁_𝑖𝑗 ∣_ 𝑌, 𝑃^(𝑝)^], the expected number of transitions from 𝑖 to 𝑗 under the current parameter estimates. For observations of stage 𝑚 followed by stage 𝑛 after 𝑡 time units (with count 𝑂_𝑚𝑛𝑡_), the expected transitions are computed by weighting all possible intermediate paths:

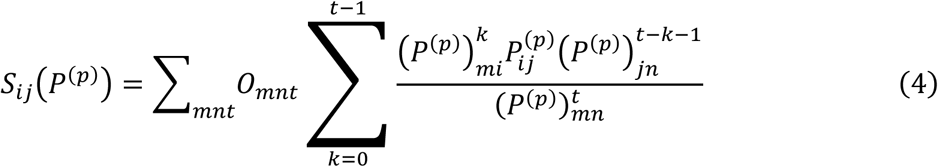

In the M-step, the transition probabilities are updated by normalizing the expected transition counts:

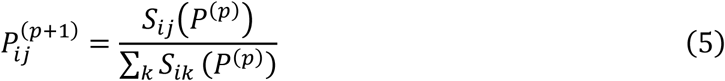

The algorithm was initialized using a naïve one-step counting estimator 𝑃^(0)^, which provides a reasonable starting point that reflects the expected structure of CKD progression, with higher probabilities of remaining in the same stage or transitioning to adjacent stages. Because the EM updates preserve structural zeros, a transition that is initialized to zero remains zero in all subsequent iterations. Specifically, if a one-step transition is not directly observed and the corresponding element of the initial transition matrix is set to zero, it receives zero expected counts during the E-step and consequently remains fixed at zero in subsequent iterations. To mitigate this locking effect, following Sherlaw-Johnson et al., the initial transition matrix was constructed without zero entries by assigning a small positive value (1 × 10^−5^) to transitions considered clinically unlikely but not impossible, rather than fixing them at zero. This initialization allows the EM algorithm to assign nonzero probability to transitions that may not be observed within a single time interval but are supported indirectly by longer-interval observations through intermediate latent paths.

The E- and M-steps were iterated until convergence, defined as the maximum absolute change in any transition probability being less than 𝜖 = 10^−5^ between successive iterations. As shown by Sherlaw-Johnson et al., this procedure yields a non-decreasing sequence of likelihood values and converges to a stationary point of the observed-data likelihood. This EM framework naturally accommodates irregular observation intervals and partially observed baseline CKD stage, avoiding the need for ad hoc imputation or exclusion of patients with incomplete data.

The EM algorithm was implemented in Python 3.7 using NumPy for matrix operations. On a standard laptop (Intel Core i7, 16 GB RAM), computation time was approximately 21 seconds when using 6-month transition intervals and 37 seconds when using 3-month transition intervals.

### 2.3 Model Validation Using Likelihood Ratio Test

We evaluated the fit of the EM-estimated transition matrix using a likelihood ratio-based comparison^44^ to assess how well the EM-based model fits the observed CKD progression data. Let 𝑛_𝑖𝑗_ denote the number of the observed transitions from CKD state 𝑖 to 𝑗, the likelihood of the observed transitions under the EM-estimated transition matrix 𝑃 is given by:

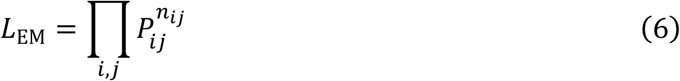

where 𝑃_𝑖𝑗_represents the estimated transition probability from state 𝑖 to state 𝑗. For reference, we also computed the likelihood based on empirical one-step transition frequencies obtained by directly normalizing observed transition counts 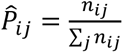 with likelihood:

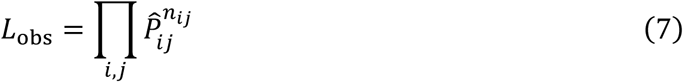

We then computed the likelihood ratio statistic as:

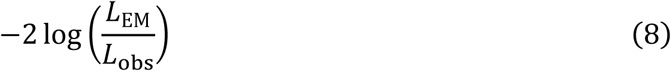

to quantify the relative improvement in fit achieved by the EM framework. Because both estimators assume the same underlying Markov structure and estimate transition probabilities over the same state space; therefore, the two models have the same number of free parameters. As a result, this statistic is not interpreted as a formal hypothesis test against a chi-squared distribution, but rather as a measure of relative model fit. Larger values of the likelihood ratio statistic indicate a greater improvement in likelihood achieved by accounting for unobserved intermediate CKD transitions through the EM framework.

## 3. Results

### 3.1 Estimated Transition Matrix

The final analytic cohort included 527 patients with small renal masses who contributed at least two eligible outpatient eGFR measurements, thereby providing at least one observable CKD transition interval. At baseline, the cohort predominantly exhibited preserved or mildly reduced kidney function, with 73.6% patients in CKD stages 1–2 and a smaller proportion in more advanced stages. This distribution is consistent with the expected kidney function profile of patients with small renal masses under active surveillance, who typically have preserved or mildly reduced kidney function at diagnosis.^45,46^

Using these longitudinal renal function data, we estimated CKD stage transition probabilities using both an empirical counting approach based on directly observed transitions and the EM-based method described above. EM-based transition matrices were estimated under two cycle lengths: a 3-month interval, selected to approximate the typical spacing between consecutive eGFR measurements, and a 6-month interval, chosen to reflect clinically relevant surveillance and imaging intervals in SRM management. The naïve transition matrix is shown in Figure 2. EM-estimated transition matrices under 3-month and 6-month cycle lengths are shown side by side in Figure 3.

**Figure 2:**
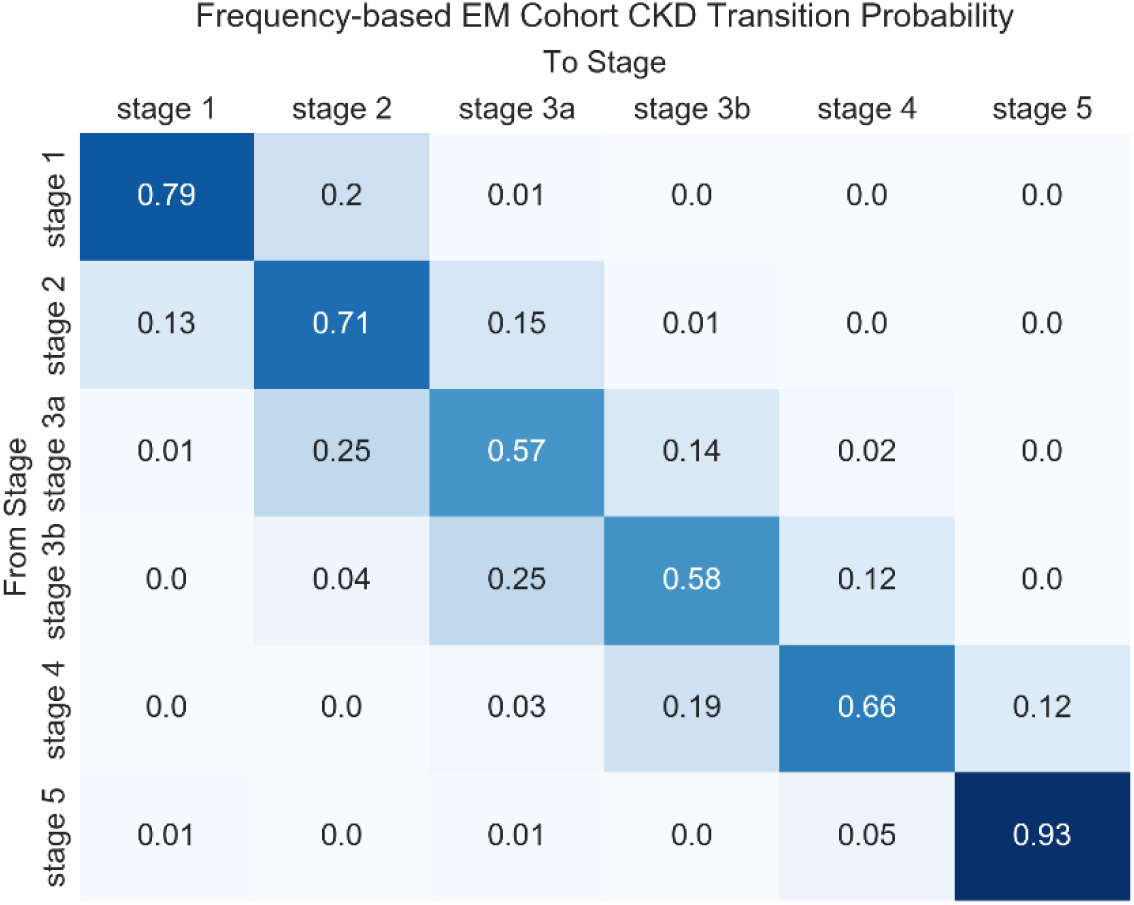
Naïve CKD Stage Transition Matrix

**Figure 3:**
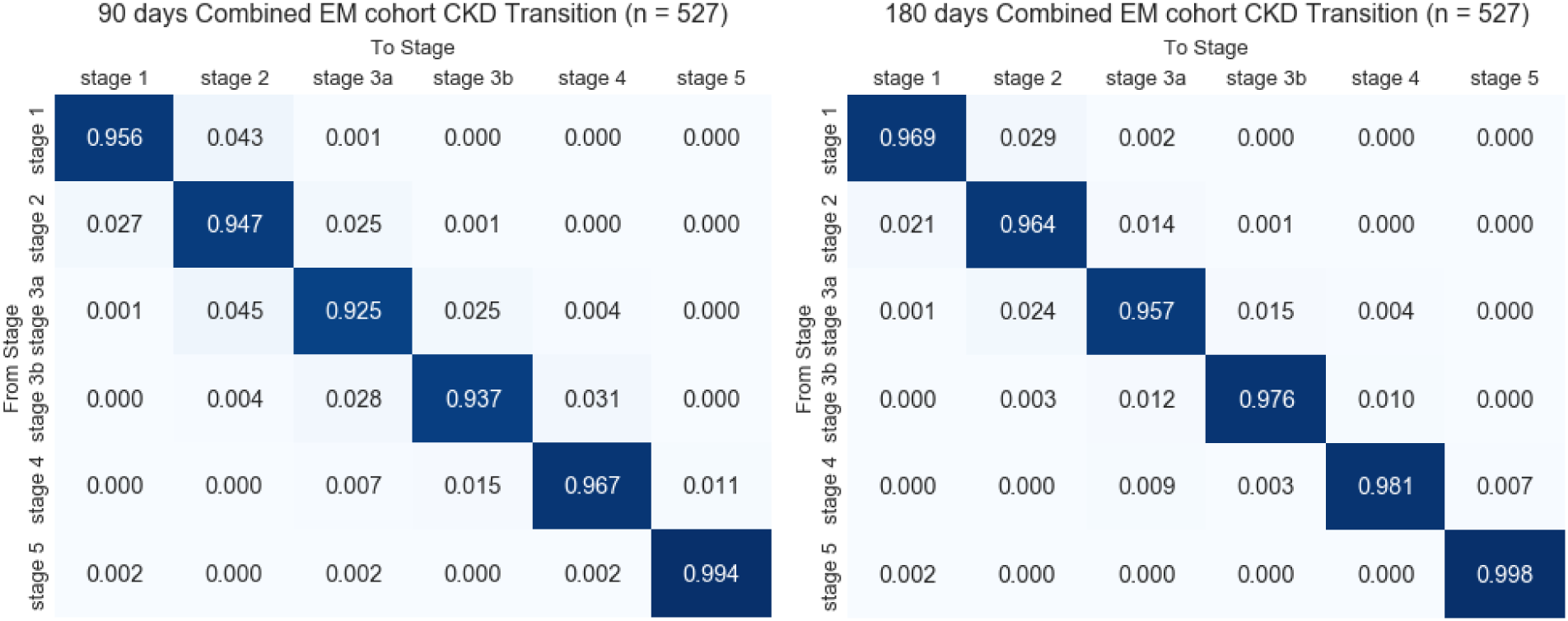
EM-Estimated CKD Stage Transition Matrices Under 3-month (left) and 6-month (right) Cycle Lengths

Under the naïve estimator, transition probabilities were derived solely from the frequency of observed transitions between consecutive CKD stages, without accounting for the time interval between observations. Although CKD progression typically reflects gradual decline in kidney function, a small number of backward stage transitions were observed in the longitudinal data. These observations were retained rather than excluded, as they reflect clinically plausible short-term changes seen in real-world practice. In some patients, transient reductions in kidney function occurred in the setting of acute illness, trauma, or other reversible conditions, followed by recovery on subsequent measurements. Patients who had received technically successful kidney transplants with subsequent ideal graft function also experienced significant improvement in CKD. Retaining these observations allowed the analysis to capture the full spectrum of observed renal function trajectories while avoiding selective exclusion of clinically meaningful events.

Because the naïve estimator treats all observed transitions equally regardless of the time between measurements, the resulting transition matrix assigns substantial probability to backward transitions (e.g., CKD stage 3a to stage 2 and stage 3b to stage 3a), reflecting short-term fluctuations in eGFR and transient changes in renal function rather than sustained improvement in underlying kidney disease. This effect is particularly pronounced in advanced CKD stages, where persistent recovery to less severe stages is relatively uncommon. By neglecting the duration of observation intervals, the naïve estimator conflates short-term variability with longer-term disease progression, leading to less stable transition patterns.

In contrast, the EM-estimated transition matrices explicitly accounted for irregular observation intervals and unobserved intermediate transitions, yielding transition patterns more consistent with expected CKD progression. Across all stages, remaining in the same CKD stage was the most likely outcome. Transition probabilities estimated under the 3-month and 6-month cycle lengths were broadly similar, demonstrating robustness of the EM approach to alternative temporal discretization. As expected, the 6-month model showed slightly higher probabilities of remaining in the same stage. The longer transition interval effectively averages short-term variability in eGFR measurements, reducing the influence of transient fluctuations and measurement noise observed over shorter intervals. Consequently, the 6-month transition matrix appears smoother and more stable than the 3-month matrix while maintaining the same overall progression structure.

Figure 4 shows EM-estimated CKD stage transition matrices stratified by age (< 65 vs ≥ 65 years) using a 6-month cycle length. In both age groups, remaining in the same CKD stage was the most likely outcome; however, patients aged 65 years or older exhibited slightly higher probabilities of forward progression to more advanced CKD stages and lower probabilities of remaining in earlier stages compared with younger patients. We also estimated EM-based transition matrices stratified by sex. As shown in Figure 5, transition patterns were largely similar between female and male patients, with dominant self-transitions and progression occurring primarily to adjacent stages. Overall, sex-related differences in transition probabilities were small, suggesting that sex had a limited impact on CKD stage transitions in this cohort.

**Figure 4:**
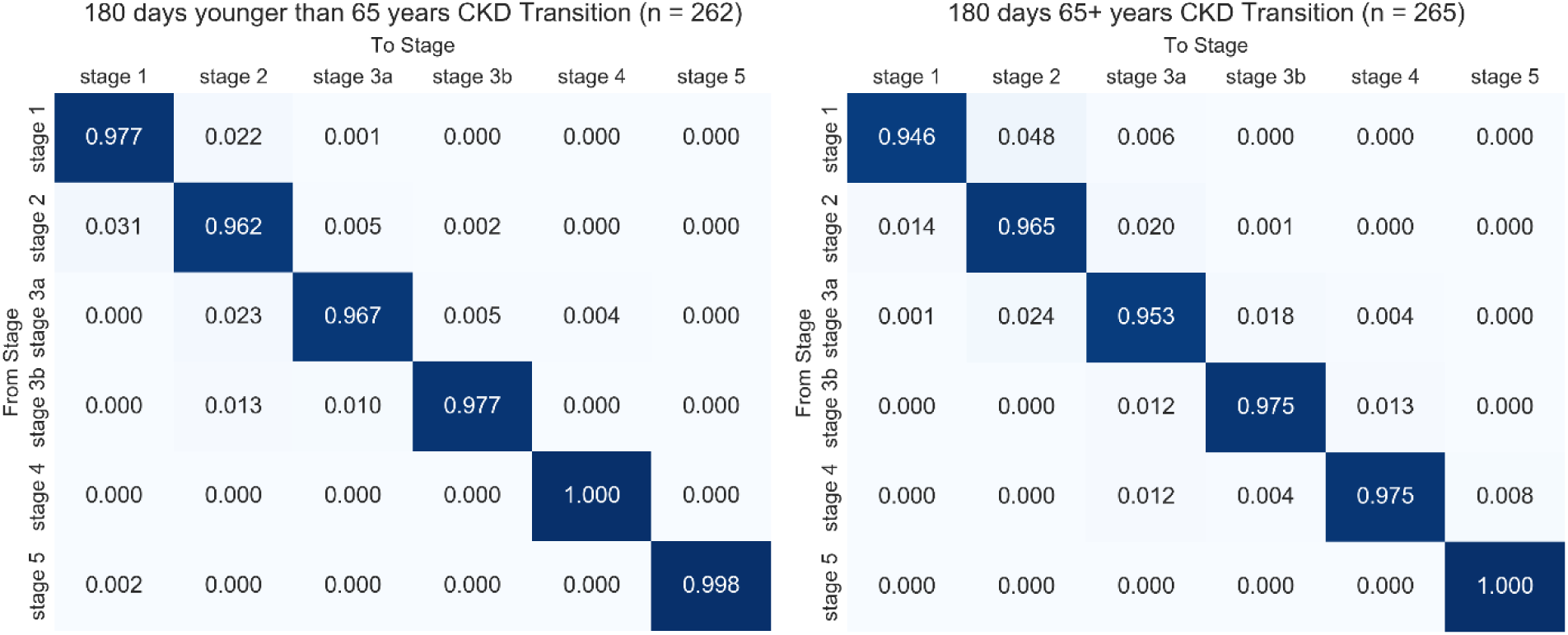
**EM-Estimated CKD Stage Transition Matrices Stratified by Age: <65 Years (left) and ≥65 Years (right) using 6-month Cycle Length**

**Figure 5:**
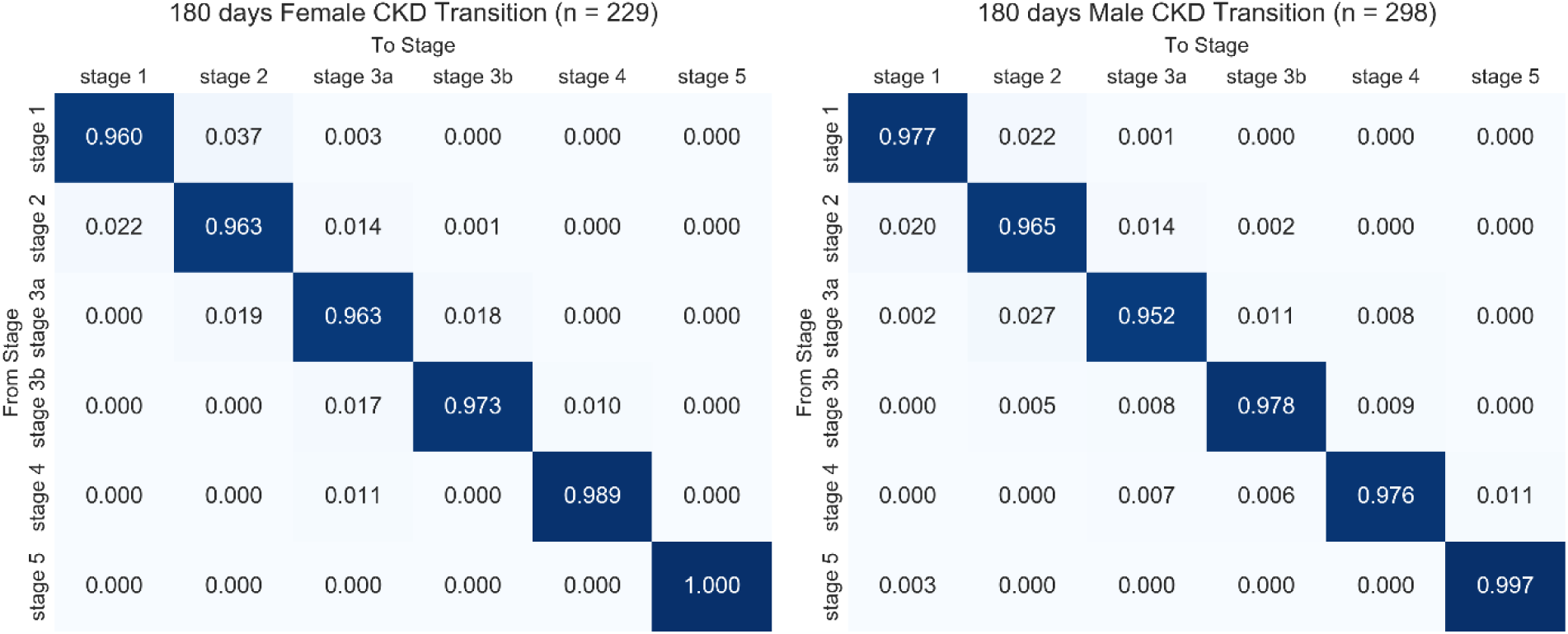
EM-Estimated CKD Stage Transition Matrices Stratified by Sex: Female (left) and Male (right) using 6-month Cycle Length

Taken together, these results indicate that the EM-based approach produces clinically plausible estimates of CKD stage progression from irregularly observed EHR data. Compared with the naïve one-step estimator, the EM framework reduces spurious backward transitions and yields consistent transition structures across alternative cycle lengths and clinically relevant subgroups. While age was associated with modest differences in the magnitude of CKD progression, the overall transition patterns were similar across age groups, and sex-related differences were minimal.

### 3.2 Model Validation Results

We evaluated the adequacy of the EM-estimated CKD transition matrix using a likelihood ratio–based measure. This validation assessed whether explicitly accounting for irregular observation intervals and unobserved intermediate transitions introduced any lack of fit relative to the empirical CKD transition structure in the data.

Across the overall cohort as well as age- and sex-stratified analyses, the likelihood ratio comparison yielded p-values of 0.999 for both the 3-month and 6-month cycle lengths, indicating no evidence that the EM-based models provided a worse fit to the observed data. Because the likelihood was computed over the same observed transition counts and state space, the resulting statistic was interpreted as a descriptive measure of model adequacy rather than a formal hypothesis test.

Overall, these results indicate that the EM-estimated transition matrices remain compatible with the empirical CKD progression patterns observed in the data while accommodating irregular follow-up intervals and unobserved intermediate transitions, both in the overall cohort and across clinically relevant subgroups.

## 4. Discussion

In this study, we estimated CKD stage transition probabilities using an EM framework applied to irregularly observed EHR data from patients with small renal masses. By explicitly accounting for interval censoring and unobserved intermediate transitions, the EM-based approach produced CKD progression patterns that were clinically plausible and consistent across alternative cycle lengths. Compared with a naïve one-step counting estimator, the EM framework reduced spurious backward transitions and yielded transition structures that better reflect the natural history of CKD. Age-stratified analyses revealed modest but systematic differences in progression patterns, whereas sex-related differences were limited, indicating that age plays a more prominent role than sex in shaping CKD progression in this cohort.

A key contribution of this work is methodological. Estimating disease progression from real-world EHR data is challenging due to irregular follow-up, interval-censored state transitions, and measurement variability. Naïve estimators that rely on observed consecutive measurements implicitly assume fixed observation intervals and may conflate short-term laboratory fluctuations with true disease progression. In contrast, the EM framework leverages information from transitions observed over varying time intervals to reconstruct the expected number of one-step transitions consistent with the observed data. This enables estimation of discrete-time transition probabilities without requiring ad hoc imputation, artificial discretization of follow-up, or exclusion of patients with incomplete observation histories. Likelihood-based validation further demonstrated that incorporating this additional structure does not distort the empirical transition patterns present in the data.

From a clinical perspective, the estimated transition matrices exhibit features that align with established understanding of CKD progression. Across analyses, remaining in the same CKD stage was the most likely outcome over a single cycle, with progression occurring primarily to adjacent stages. Although backward transitions were observed, they occurred infrequently and were consistent with known short-term variability in renal function, particularly in outpatient settings. Inpatient measurements were excluded to minimize acute illness–related fluctuations; however, backward stage transitions were still observed and were intentionally retained, as they reflect real-world recovery of renal function following severe illness or trauma, reflective of true clinical environment rather than data error. Retaining these observations allowed the model to capture the full spectrum of outpatient renal function dynamics observed in practice. Age-related differences in progression were consistent with known declines in renal function at older ages, whereas sex-related variation appeared comparatively limited. Together, these findings suggest that the EM-based estimates capture clinically meaningful aspects of CKD progression while remaining grounded in observed EHR data.

In this study, mortality was modeled separately rather than as a direct absorbing state within the CKD transition framework. This decision was informed by both cohort characteristics and data considerations. Patients with small renal masses are often managed under surveillance and typically present with early-stage CKD, for whom short-term kidney-related mortality risk is low. In addition, mortality and cause-of-death information were incomplete for a subset of patients. Modeling mortality separately enabled more stable estimation of stage-to-stage renal function dynamics while avoiding overfitting sparse stage-to-death transitions. In downstream decision models, mortality can be incorporated as a competing-risk process conditional on CKD stage, providing greater flexibility for evaluating long-term clinical outcomes.

We included a two-year pre-diagnosis lookback period to better characterize underlying CKD trajectories among patients with SRMs. Although CKD progression could be defined starting at the time of diagnosis to establish a strictly aligned temporal origin, SRMs are frequently detected incidentally, and tumor presence often precedes clinical discovery by an unknown duration.

Consequently, renal function dynamics relevant to SRM management may already be evolving prior to the recorded diagnosis date. In addition, many patients, particularly those undergoing early treatment, had limited outpatient eGFR measurements between diagnosis and first intervention, such that restricting the analysis to post-diagnosis observations would substantially reduce available transition information and fail to capture early CKD dynamics. Incorporating a two-year lookback period allowed recovery of these otherwise unobserved transitions and improved stability of transition probability estimation while maintaining clinical relevance to patients entering SRM surveillance. Nevertheless, inclusion of pre-diagnosis measurements may introduce temporal misalignment between disease detection and modeled progression, and this potential source of bias should be considered when interpreting the estimated transition probabilities.

Several limitations of this study should be acknowledged. First, the transition matrices were estimated for an all-comers cohort to characterize average CKD progression patterns in patients with SRMs. While this population-level approach is appropriate for estimating general transition probabilities, CKD progression is known to vary by comorbidity burden, baseline risk factors, and treatment history. If data availability permits, future work could extend this framework to more granular subgroup analyses, such as stratification by diabetes status, cardiovascular comorbidities, or other clinical risk factors.

Second, the EM framework estimates discrete-time transition probabilities by maximizing the observed-data likelihood and does not explicitly model the rate of progression between CKD stages. Survival models or CTMC formulations may be better suited for estimating transition intensities or hazards. However, such approaches typically require stronger assumptions regarding time homogeneity or parametric hazard structures and are often more computationally intensive. In contrast, the discrete-time EM approach provides mathematically simple and computationally efficient transition probability estimates that are well suited for downstream decision-analytic models. Future work could directly compare EM-based estimates with those obtained from continuous-time or hidden Markov model formulations to assess consistency across modeling paradigms.

Third, the analysis relied on outpatient EHR data available within a single health system and therefore reflects the information captured during routine clinical care rather than complete longitudinal patient histories. Although inpatient measurements were excluded to reduce acute illness–related fluctuations, renal function changes occurring during hospitalizations or care received outside the health system may not have been fully observed. As a result, some transitions may appear irregular or less stable due to incomplete clinical information rather than true biological variability. This limitation is inherent to real-world EHR data and reflects common challenges encountered when modeling disease progression using routinely collected clinical records.

Additional limitations include potential misclassification of CKD stage due to biological and measurement variability in eGFR, particularly near stage boundaries where small fluctuations in laboratory values may result in shifts between adjacent CKD categories. Although the EM framework mitigates some effects of irregular observation and short-term fluctuation, residual measurement noise may still influence estimated transitions. Lastly, model validation focused on internal consistency with observed transition data; external validation in independent cohorts remains an important direction for future research.

Despite these limitations, the proposed framework has important implications for decision modeling in kidney cancer and other chronic disease contexts. The EM-estimated transition matrices provide a principled way to translate irregular EHR data into discrete-time progression models that can be directly embedded within Markov decision processes, microsimulation models, and cost-effectiveness analyses. In the context of small renal mass management, accurate modeling of CKD progression is critical for evaluating trade-offs between cancer control and long-term kidney function preservation. More broadly, this approach offers a generalizable pathway for leveraging real-world data to inform evidence-based, patient-centered decision support in settings characterized by heterogeneous follow-up and latent disease progression.

## 5. Conclusion

In conclusion, the Expectation–Maximization approach enabled estimation of clinically plausible CKD stage transition probabilities from irregularly observed EHR data in patients with small renal masses. Compared with naïve methods, the EM framework reduced spurious backward transitions and produced stable progression patterns across alternative cycle lengths and subgroup analyses. These results provide reliable, model-ready inputs for future decision-analytic studies of SRM management and demonstrate a practical framework for leveraging real-world clinical data to inform disease progression modeling under irregular follow-up.

## Data Availability

The data used in this study were obtained from the University of Virginia Small Renal Mass registry. Due to institutional and ethical restrictions, these data are not publicly available but may be made available from the corresponding author upon reasonable request and with appropriate institutional approvals.

